# Attention and memory in Parkinson’s disease: a discriminant analysis approach

**DOI:** 10.64898/2026.06.17.26355843

**Authors:** Marco Calabria, Lucas Guallar, Carmen García-Sánchez, Berta Pascual Sedano, Jaime Kulisevsky

## Abstract

**Background:** Cognitive impairment in Parkinson’s disease (PD) is highly prevalent and heterogeneous. Assessing multiple cognitive domains is challenging and risks redundancy. This study evaluated whether a discriminant analysis approach could optimize the selection of specific tasks and measures for identifying attention and memory deficits in PD.

**Methods:** Thirty PD patients and 25 cognitively unimpaired (CU) controls completed four experimental tasks: two assessing attention (flanker and spatial Stroop), one for recognition memory, one for working memory (n-back). Following group-level difference analyses, a discriminant analysis was performed to identify which tasks, and performance metrics possessed the highest sensitivity for distinguishing PD patients from CU individuals.

**Results:** At the group level, PD patients exhibited significantly worse conflict costs in both attention tasks and lower sensitivity scores (d’) in the recognition memory task compared to CU controls. The discriminant analysis revealed that time-based measures from the spatial Stroop task and the sensitivity score from the recognition memory task provided the highest discriminating power to differentiate between the two groups.

**Conclusion:** These findings suggest that cognitive deficits in PD can be identified with high diagnostic accuracy using a targeted subset of metrics, eliminating the need for extensive and redundant neuropsychological testing batteries for attention and memory, without needing an extensive number of cognitive tasks for attention and memory.

## 1. Introduction

Cognitive impairment is a highly prevalent and disabling non-motor feature of Parkinson’s disease (PD) that may emerge early in the course of the illness [1–5]. In recognition of its clinical relevance, formal diagnostic criteria for Parkinson’s disease–mild cognitive impairment (PD-MCI) have been established [6], recommending the presence of either cognitive decline on a brief cognitive assessment or impairment on at least two neuropsychological tests within a more comprehensive evaluation. However, because the cognitive profile of PD is notably heterogeneous [7], clinicians and researchers require reliable, sensitive measures to differentiate between cognitively unimpaired and impaired individuals [8].

While substantial progress has been made, it is well-established that specific cognitive domains, namely executive function and attention, are vulnerable even in the early stages of PD [9–12]. Recently published clinical test recommendations for attention, working memory, and executive abilities provide invaluable diagnostic guidelines for clinicians tracking cognitive decline [13]. Nonetheless, given the wide variability of cognitive phenotypes within the PD population [7], a persistent challenge remains: identifying reliable, highly discriminative measures that can isolate cognitive impairment without requiring exhaustive testing of unaffected domains.

In line with this, recent research suggests that traditional tests spanning multiple domains may actually reflect a unidimensional structure, compatible with a common, underlying deficit [14]. This underscores the need to target cognitive decline in PD more precisely by using a streamlined, targeted battery of tests to reduce redundancy and improve the validity of cognitive assessment.

Given this context, the present study investigates the detection of cognitive deficits in PD patients, focusing on identifying the most discriminant tasks and performance measures. We complemented standard clinical guidelines with computer-based tasks, which were adapted from a previous study on patients with mild cognitive impairment and dementia [15]. To achieve this, we first evaluated the performance of PD patients on these tasks for memory and attention. We then employed a discriminant analysis to identify the specific performance metrics that most effectively distinguish between cognitively impaired and unimpaired individuals.

Attentional and executive dysfunctions in PD are widely considered hallmark features of the disease [9–12]. These deficits are primarily attributed to dopamine-dependent dysfunction within frontostriatal networks [3], are affected to different degrees and at different stages of PD, and potentially associated with behavioural and emotional symptoms [10]. Studies using interference paradigms, such as the Stroop and flanker tasks, have shown that patients with PD are impaired due to deficits in resolving attentional conflict, maintaining vigilance, and inhibiting inappropriate responses [e.g., 12]. .

Building on this literature, in our study we employed two interference tasks that are thought to assess different levels of conflict processing: stimulus-level interference (flanker task [5]) and response-level interference (spatial Stroop task [6]). In the context of PD, findings from flanker task studies suggest that the magnitude of the interference effect produced by incongruent flankers varies as a function of disease duration and dopaminergic medication status .[16]. Studies reporting greater interference effects in patients with PD compared to cognitively unimpaired controls often interpret these findings as reflecting an impaired ability to suppress competing response tendencies .[17]. Such deficits have also been observed under conditions with longer inter-trial intervals, suggesting that response inhibition deficits remain persistent throughout the task [18].

In contrast, although the verbal color-word Stroop task is considered a gold-standard clinical measure of executive functioning in PD [13], relatively few studies have examined the spatial Stroop version. Nevertheless, given the well-documented impairments in visuospatial processing and attention in PD [19], greater interference effects in patients than in cognitively unimpaired controls would be expected under conflict conditions. In the spatial Stroop task, interference arises when the direction indicated by an arrow is incongruent with its spatial location on the screen [20]. This manipulation closely resembles that used in the Simon task, which assesses response–spatial incompatibility. Studies using the Simon task have shown that individuals with PD often exhibit increased interference effects, which have been attributed to deficits in response inhibition [16].

Alongside attentional deficits, long-term memory decline is among the most prominent cognitive complaints in the early stages of PD [21,22]. PD pathology directly affects the hippocampus [23,24], but also striatum and amygdala [25], leading to impairments in recollection processes while relatively preserving familiarity-based recognition, at least at early stages of the diseases. Although memory deficits in PD were traditionally considered to reflect retrieval failures secondary to frontal-executive dysfunction, more recent evidence has challenged this view, reporting impairments in the encoding and consolidation of new information [26].

Based on this evidence, we included a face recognition memory task to assess non-verbal long-term memory performance. In this task, participants first encoded a series of unfamiliar faces and were subsequently required to identify them among a larger set of faces comprising both previously presented and novel stimuli. This paradigm allows the estimation of signal detection measures, including sensitivity and response bias, thereby providing a more nuanced assessment of recognition memory performance than traditional accuracy measures alone.

To further characterize memory functioning, we also included a working memory task, which assesses the short-term maintenance and manipulation of information. Working memory deficits are consistently reported in PD and are thought to reflect dysfunction within frontoparietal and frontostriatal networks [27]. Specifically, we employed an n-back task in which participants were required to determine whether the current stimulus (a letter) matched the one presented n trials earlier. Like the face recognition task, performance can be quantified using measures of sensitivity and response bias. The n-back paradigm is widely used to assess working memory capacity in both healthy and clinical populations. In PD, poorer performance on n-back tasks has been associated not only with working memory impairments but also with broader deficits in processing speed [28]. Furthermore, performance appears to be modulated by dopaminergic medication status, with differences reported between patients assessed in the ‘on’ and ‘off’ medication states [29].

Therefore, based on the previous literature and the combination of experimental tasks employed in the present study, we hypothesized that, at the group level, patients with PD would show lower performance across most cognitive tasks compared with cognitively unimpaired individuals without PD. More specifically, we expected the most consistent group differences to emerge in tasks assessing attention and working memory, as these functions are closely linked to executive processes, the cognitive domain most frequently impaired in PD. The two attention tasks were designed to assess executive control under different forms of conflict. The flanker task measures interference arising at the stimulus level, whereas the spatial Stroop task assesses interference at the response level. Both tasks require the inhibition of irrelevant information and the resolution of competing response tendencies. Similarly, working memory is considered a core component of the executive control system [30], and therefore we expected patients with PD to show difficulties updating and maintaining information in the n-back task.

In contrast, we anticipated more modest impairments in the recognition memory task. Because our sample consisted of individuals without dementia, and recognition memory is generally considered relatively preserved during the early stages of PD, group differences were expected to be smaller than those observed for attention and working memory measures.

Finally, discriminant analyses were conducted to identify which tasks and specific performance measures were most sensitive to cognitive impairment. For the attention tasks, both reaction time and accuracy measures were considered, whereas for the memory tasks we examined signal detection indices, including sensitivity index, which reflects the ability to discriminate target from non-target stimuli, and response bias index, which reflects the decision criterion adopted when making judgments under uncertainty.

## 2. Methods

### 2.1. Participants

Participants with PD were recruited from the Movement Disorders Unit at the Hospital Sant Pau in Barcelona (Spain) based on the following criteria: a) confirmed diagnosis of PD according to international criteria [31] by a neurologist specialized in movement disorders; b) aged between 55 and 75 years; c) exhibiting severity ranging from mild to moderate (Hoen-Yahr stage 2-3); and d) availability to participate on a voluntary basis. Exclusion criteria for patients included: a) the presence of atypical parkinsonism; b) Montreal Cognitive Assessment (MoCA) score < 21/30, used to exclude cases of PD dementia according to normative data for the Spanish population [32] ; c) the presence of comorbidities, sequelae, or any disorder that could interfere with a diagnosis of PD; and d) any condition that could interfere with the performance of computerized cognitive tasks. All patients were receiving L-Dopa medication throughout the study, and none were in an “off” state during the assessment sessions.

During the selection process, 33 participants were initially recruited. Of these, three were excluded due to a MoCA score < 21/30, resulting in a final PD sample of 30 participants (see Table 1).

**Table 1.** Sociodemographic and clinic characteristics of the sample.

|  | **PD patients**<br>**(n=30)** | **CU individuals**<br>**(n=25)** | **p values** |
| --- | --- | --- | --- |
| Age (years) | 66.8 (6.4) | 68.7 (3.8) | .205 |
| Sex (Female/Male) | 12/18 | 11/14 | .764 |
| Education (years) | 15.2 (4.1) | 13.7 (2.6) | .123 |
| MoCA score | 23.9 (2.1) | 28.5 (.9) | < .001 |
| Disease duration (months) | 81.6 (36.3) | - |  |

Cognitively unimpaired (CU) individuals were recruited through the Universitat Oberta de Catalunya (UOC) from a pool of participants who had previously taken part in studies on aging and cognition. Inclusion criteria were: (a) age between 55 and 75 years; and (b) availability to participate on a voluntary basis. Exclusion criteria included: (a) a known history of neurological or psychiatric disorders, or any condition that could interfere with the performance of computerized cognitive tasks; and (b) a Montreal Cognitive Assessment (MoCA) score < 21/30, used to exclude cases of cognitive impairment according to normative data for the Spanish population [32]. Based on these inclusion and exclusion criteria, a total of 25 participants were selected.

### 2.3. Tasks and procedure

Before starting the test session, participants provided written informed consent after receiving a full explanation of the study. Additionally, the MoCA was administered to obtain a recent assessment relevant to the inclusion criteria regarding cognitive decline. All clinical information was retrieved from the patients’ hospital medical records. The study protocol was approved by the ethics committee at the UOC (ref. CE22-RC01), and participants were given an information sheet detailing the study protocol along with contact information for any further questions after the session.

se These tasks are freely available at https://osf.io/fgu9q/files/osfstorage. The tasks were adapted from those previously employed in studies involving individuals with neurodegenerative diseases, including mild cognitive impairment and Alzheimer’s disease [15]. The tasks were implemented in DMDX [33], which was also used to present the stimuli and collect responses. Here, we provide a detailed description of the tasks.

*Flanker task.* Participants view a horizontal array composed of five black arrows, each pointing either to the left or to the right. The central arrow represents the target stimulus, while the flanking arrows act as distractors. Participants are asked to indicate, as quickly and accurately as possible, the direction of the central arrow by pressing one of two designated response keys, one for left and one for right. Two types of trials are presented: congruent, when all arrows point in the same direction, and incongruent, when the flanking arrows point opposite to the target. The task consists of two blocks of 48 trials (96 in total), with 75% congruent (n = 72) and 25% incongruent (n = 24) trials. The structure of the trial is the following: (a) a fixation cross displayed for 500 ms, followed by (b) the stimulus array, which remains visible until a response is made or for a maximum duration of 2000 ms.

*Spatial Stroop task.* In this paradigm, a single arrow is presented on either the left or right side of the display. The task manipulates spatial correspondence between the arrow’s direction and its on-screen position. In congruent trials, the arrow points toward the same side as its location, whereas in incongruent trials, the arrow’s direction and position are opposite. Congruent and incongruent trials occur with equal frequency (50% each). The experiment includes 192 trials organized into four blocks of 48. Participants indicate the direction of the arrow by pressing one of two response keys on the keyboard (e.g., one or left and one for right). The structure of the trial is the following: (a) central fixation cross presented for 500 ms, followed by (b) the presentation of the target arrow, which remains visible until a response is made or for a maximum of 2000 ms.

*Face recognition memory task.* The stimuli consisted of 48 grayscale photographs of unfamiliar faces (24 male, 24 female), obtained from publicly available image databases and online sources and displayed against a uniform gray background. The task has two phases, such as encoding and recognition. During the encoding phase, participants viewed half of the faces, each presented twice, and performed a sex classification task to promote deeper encoding and maintain attention. Each trial began with a central fixation cross shown for 500 ms, followed by a face presented for 3000 ms. Participants indicate whether the face was male or female by pressing one of two response keys and they are required to memorize the faces.

After a 10-minute interval, participants complete the recognition phase, which includes all previously shown faces (old items) along with 24 novel faces (new items). The structure of the trial is the following: (a) a fixation cross (500 ms), followed by (b) a face appearing and remaining visible until a response is made or for a maximum of 5,000 ms. Participants are asked to judge whether each face had been seen earlier by pressing one of two keys.

*N-back task.* In this sequential letter-matching task, individual letters are presented one at a time, and participants indicate whether each letter matches the one shown on the immediately preceding trial. The task consists of four blocks, each containing 25 trials, with 7 target trials (28%) and 18 non-target trials (72%). On each trial, participants respond using one of two keys to signal a match or non-match. Trials follow a consistent structure: (a) a fixation cross is presented at the center of the screen for 500 ms, followed by (b) the target letter, which remains visible for up to 1500 ms or until a response is made.

The task administration always began with the recognition memory task to allow sufficient time between the encoding and retrieval phases. Between these two phases, attentional tasks were administered in a random order, with the final task being the n-back task.

The total duration of each session was a maximum of one hour, including the explanation of the study, signing of the consent form, administration of the MoCA, the experimental tasks, and a final debriefing with the participants.

## 3. Data analysis

For the *Flanker and Stroop tasks*, RTs and accuracy were analysed using mixed-effects models implemented with the lme4 package. Log-transformed RTs were analysed using linear mixed-effects models, while accuracy data were analysed using logistic mixed-effects models (binomial family). Fixed effects included Group (Control vs. PD) and Congruency (Congruent vs. Incongruent), and random intercepts for participants and items were included in the accuracy models. Standardized covariates (MoCA, age, and education) were additionally included to control for individual variability in global cognitive status, age, and years of education.

For the *memory and n-back tasks*, sensitivity (d′) and response bias (c) were computed according to signal detection theory: d′ = Z(Hit Rate) − Z(False Alarm Rate) and c = −½[Z(Hit Rate) + Z(False Alarm Rate)]. A multiple linear regression analysis (*lm*) was conducted with d′ and c as dependent variables, and Group (Control vs. PD), age, education, and MoCA scores as independent variables.

Model assumptions were evaluated by inspecting residual distributions, leverage, and outliers using the performance and see packages. Multicollinearity was assessed with variance inflation factors (VIF) using the car package.

The detailed information of each model is reported in Appendix A - Supplementary Material.

For the *canonical discriminant analysis*, a multivariate linear model was first fitted to explore overall group differences across cognitive measures. Canonical discriminant analysis (implemented with the *candisc* package) was then applied to identify the linear combinations of variables that maximally separated the groups, providing an initial assessment of multivariate group structure.

Subsequently, to identify the most predictive cognitive markers for group classification, a Least Absolute Shrinkage and Selection Operator (LASSO) logistic regression was performed using the *glmnet* package. All continuous predictors were standardized to ensure the regularization penalty was applied uniformly across variables with different measurement scales. A 10-fold cross-validation procedure was employed to ensure generalizability and prevent overfitting, and the optimal regularization parameter was determined based on the minimum binomial deviance.

The data associated with this study have been made publicly available via the Open Science Framework (OSF) and can be accessed at https://doi.org/10.17605/OSF.IO/VGHY8.

## 4. Results

### 4.1. Cognitive outcomes

***Flanker task.*** For accuracy, the model revealed a significant main effect of Congruency (D = - 1.69, z = −4.51, p < .001), indicating that participants were less accurate in incongruent trials (0.97) compared to congruent trials (0.99). In contrast, the main effect of Group was not significant (D= - 0.36, z = −0.53, p = .59), and the Congruency × Group interaction was also non-significant (D = −0.44, z = −0.93, p = .35) (see Figure 1).

**Figure 1.**
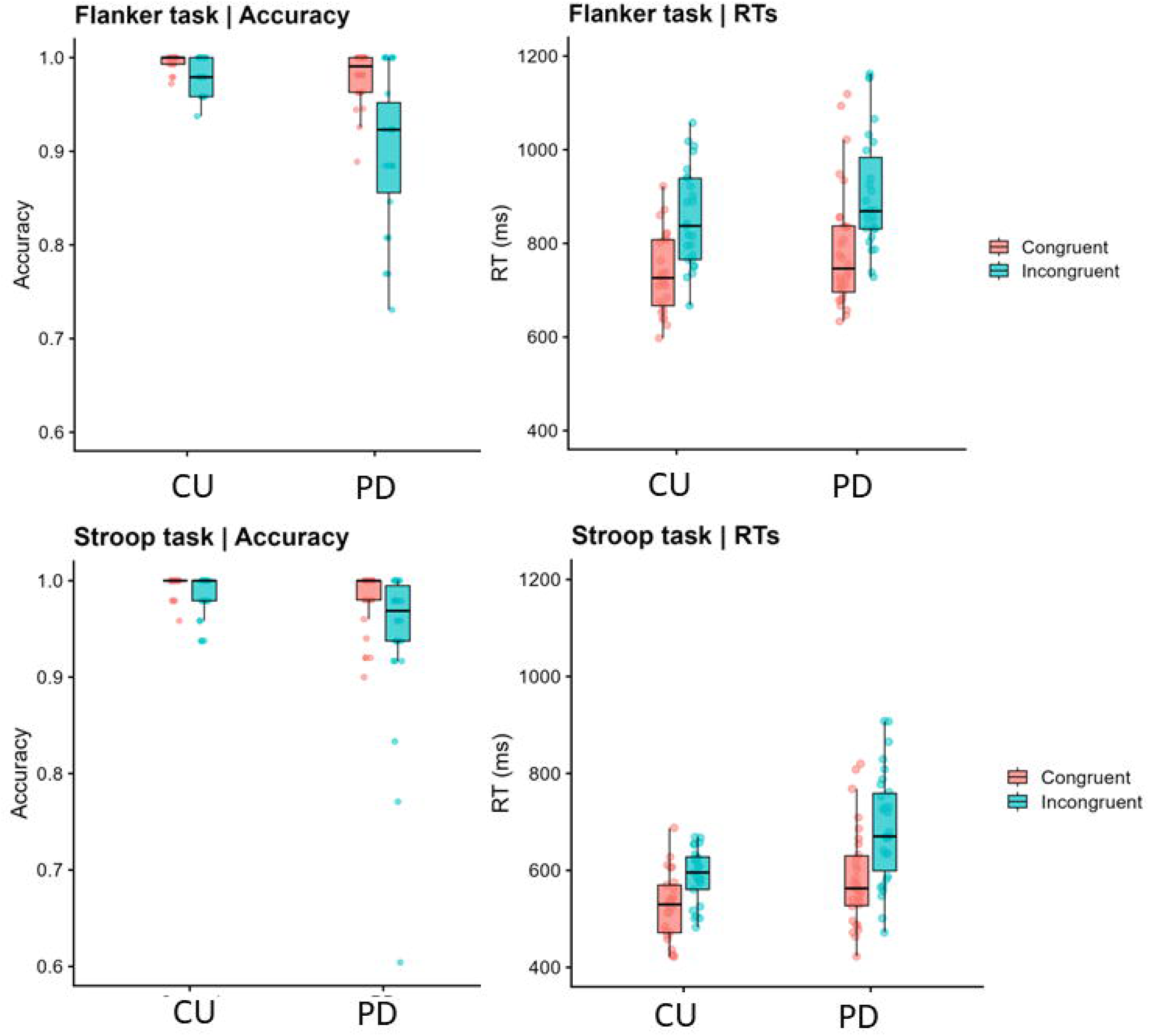
Accuracy and reaction times in PD patients and cognitively unimpaired individuals in the flanker and spatial Stroop tasks.

For RTs, the model revealed a significant main effect of Congruency (D = 0.15, t(32.3) = 8.70, p < .001), suggesting that incongruent trials (867 ms) resulted in significantly longer RTs than congruent trials (731 ms). While there was no significant main effect of Group (D = 0.017, p = .81), a significant Congruency × Group interaction was observed (D = 0.03, t(2712) = 2.46, p = .014). This interaction indicates that the conflict effect was significantly larger in the PD group (152 ms) compared to the CU group (120 ms).

***Stroop task.*** For accuracy, the model revealed a significant main effect of Congruency (D = − 1.28, z = −2.30, p = .022), where participants were significantly less accurate on incongruent trials (0.95) compared to congruent trials (0.98). There was a marginal main effect of Group (D = −1.52, z = −1.81, p = .071), suggesting a trend toward lower overall accuracy PD group compared to CU individuals. In contrast, the Congruency × Group interaction was not significant (D = −0.11, z = −0.20, p = .841).

For RTs, the model revealed a significant main effect of Congruency (D = 0.12, t(8.3) = 7.09, p < .001), incongruent trials (623 ms) showed longer RTs than congruent trials (542 ms). While the main effect of Group was not significant (D = 0.03, p = .69), we found a significant Congruency × Group interaction (D = 0.04, t(5133) = 3.67, p < .001). This interaction demonstrates that the Stroop interference effect was significantly larger in the PD group (95 ms) compared to the CU group (67 ms).

***Memory task.*** For d’ values, the analysis revealed a significant main effect of Group (D = - 1.08, t(50) = −3.24, p = .002), indicating that the PD group (0.27) had significantly lower sensitivity scores compared to the CU group (1.35).

For c values, there was no significant effect of Group (D = 0.14, t(50) = 0.67, p = .508), suggesting that response bias scores did not differ significantly between PD (0.40) and CU individuals (0.26).

***N-back task.*** For d’ values, the analysis revealed a non-significant effect of Group (D = −0.68, t(50) = −1.63, p = .109), indicating that sensitivity scores did not differ significantly between the PD group (0.94) and CU individuals (1.62) (see Figure 2).

**Figure 2.**
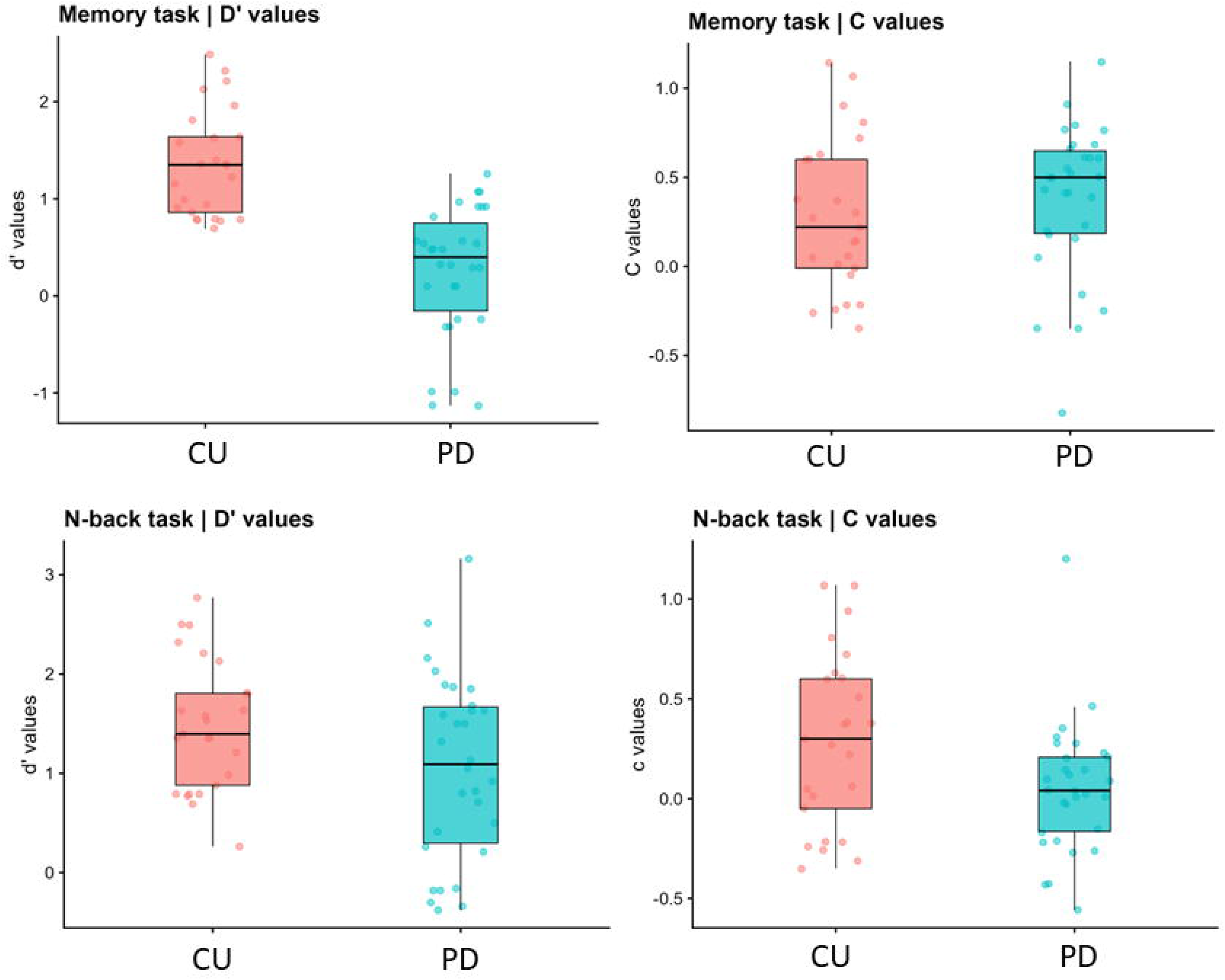
D’ and c values in PD patients and cognitively unimpaired individuals in the recognition memory task and n-back task.

For c values, the analysis revealed a non-significant effect of Group (D = −0.09, t(50) = −0.44, p = .663), indicating that response bias scores did not differ significantly between PD (0.12) and CU individuals (0.21).

### 4.2. Discriminant analysis

The model yielded a single canonical function that was highly effective in separating the groups, with a canonical correlation of .807 (CanRsq = .652) and the class means (centroids) for the two groups were −1.47 and 1.22, indicating a clear spatial separation along the canonical dimension.

After performing LASSO, four key variables were selected that best characterize the difference between groups, with the following standardized coefficients: a) d’ values from the memory task (D = −1.63), b) RTs for incongruent trials in the Stroop task (D = 0.72), c) RTs for the congruent trials in the Stroop task (D = −0.54), and d) the interference cost in the Stroop task (D = 0.14).

The model demonstrated high diagnostic accuracy, correctly classifying 92.7% of the participants (95% CI [0.82, 0.98]), with a Kappa statistic of 0.85, indicating strong substantial agreement. The model achieved high balanced performance, with a sensitivity of 93.3% and a specificity of 92.0%. The cross-validated LASSO model results reinforce the findings with an overall accuracy of 85.5% (95% CI [0.73, 0.94]) with a sensitivity of 90.0% and as specificity of 80.0% (see Figure 3).

**Figure 3.**
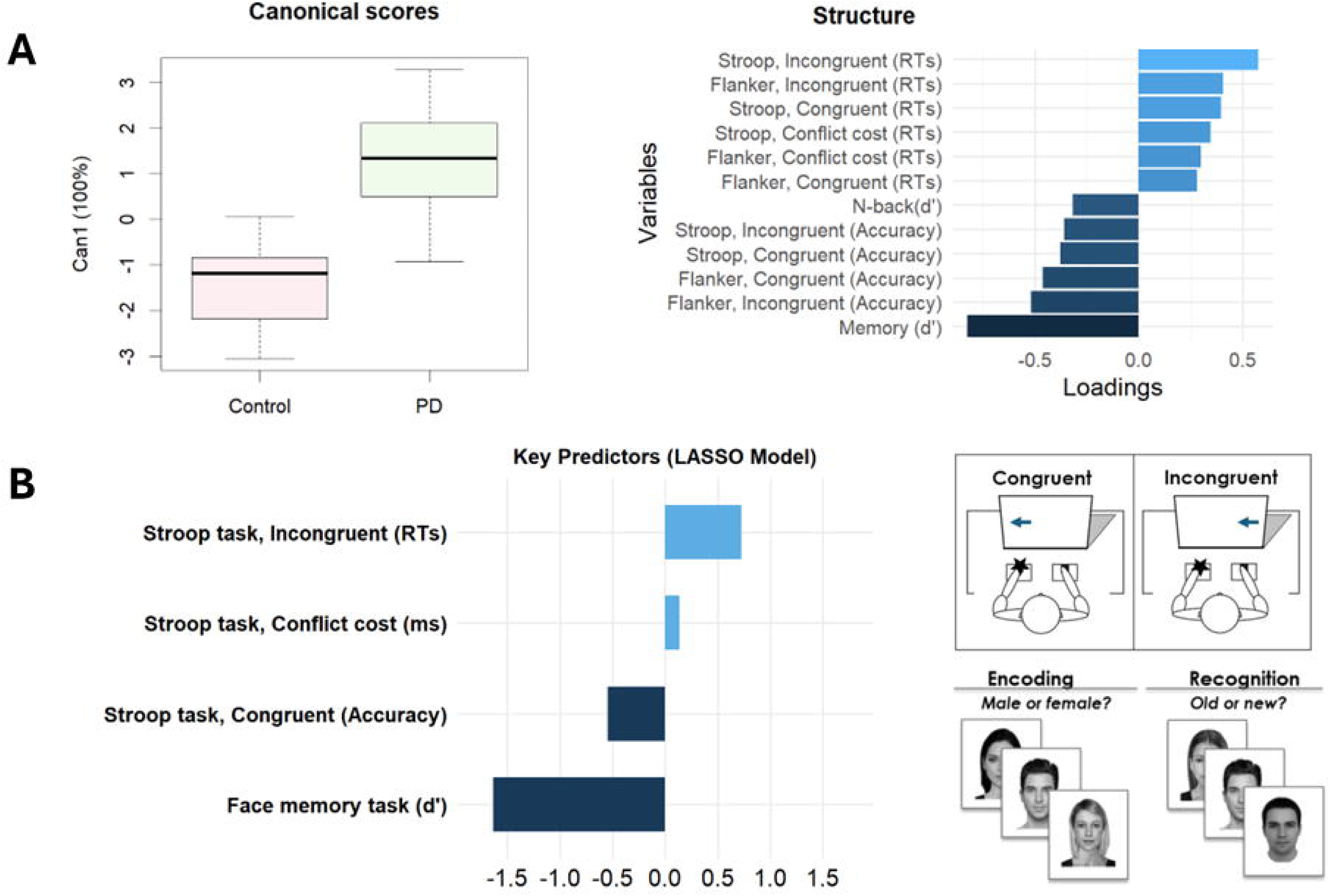
Results of the discriminant analysis: canonical scores, structure, and key predictors from the LASSO model.

## 5. Discussion

In the present study, we investigated the presence of attention and memory deficits in patients with PD using four experimental tasks. To this end, we first evaluated behavioural performance across these tasks and subsequently conducted a discriminant analysis to identify which specific tasks and performance metrics could effectively differentiate PD patients from CU individuals.

Regarding cognitive performance, our findings revealed that the two groups exhibited distinct performance profiles depending on the task type and cognitive domain. The most consistent results emerged within the attention domain; across both attentional tasks, PD patients exhibited larger conflict costs than CU individuals. This indicates a specific deficit in managing response efficiency when confronted with incongruent information at both the stimulus and response levels. Crucially, this conflict cost in RTs was accompanied by a non-significant group difference in overall processing speed, as well as highly similar accuracy rates—with the sole exception of a marginally significant group difference in the Stroop task. This pattern suggests that the observed deficits are not explained by generalized cognitive or motor slowing, but rather by a specific reduction in the efficiency of the attentional network.

While we initially predicted a stronger impairment in the Stroop task compared to the flanker task, our results suggest that conflict or interference suppression deficits in PD may not be tied strictly to the level of motor response inhibition. Instead, these deficits appear to involve the general resolution of conflicting information. Furthermore, conflict monitoring seems to remain preserved in this cohort, given that overall processing speed did not differ between PD patients and CU individuals.

Our initial hypothesis—that interference suppression deficits would be more pronounced in the Stroop task—was based on the rationale that interference suppression would be disproportionately affected by the motor demands inherent to PD. However, our findings align with literature demonstrating that the specific nature of the interference suppression or response inhibition task may be less critical, as the magnitude of conflict effects is often comparable across different paradigms [16], even when comparing patients in “on” versus “off” medication states. While evaluating medication states was beyond the scope of this study—as all included patients were tested in the “on” state—our results are consistent with previous studies utilizing the flanker task to measure interference suppression and attention in this clinical population [17,18].

Regarding the memory domain, we found that patients with Parkinson’s disease (PD) exhibited poorer performance only in the recognition memory task, while their performance on the task measuring working memory efficiency remained intact. Hence, these results align with previous research reporting recognition memory deficits in PD patients during both encoding and retrieval, despite mixed findings regarding the dissociation between familiarity and recollection—processes primarily dependent on the hippocampus [23,25]. Furthermore, our findings suggest that while PD patients suffered a genuine impairment in memory sensitivity, their decision-making strategy remained preserved. This indicates that although their underlying memory trace or retrieval capability was degraded, their strategic control over decision criteria during recognition was maintained.

Contrary to our expectations, patients with PD did not exhibit working memory impairments, at least based on their performance in the 1-back task. We had predicted that PD patients would show deficits, given that the n-back is a standard measure of working memory and executive functioning [30] domains typically affected in this population. One possible explanation for this unexpected finding is that the task load was not sufficiently demanding, as it required holding only a single item in memory. Alternatively, this outcome may have been modulated by medication status, as working memory deficits in PD have been shown to depend on the daily levodopa equivalent dose [27].

Interestingly, the results of the discriminant analysis align closely with the group comparisons of cognitive outcomes, helping to identify the specific tasks and performance measures that best differentiate the two groups. The selected variables yielded high diagnostic accuracy, demonstrating both high sensitivity (93.3%) and specificity (92.0%). Specifically, the model identified the recognition memory task, alongside the spatial Stroop task, as the primary measures discriminating between CU individuals and PD patients. The finding that the attention domain was characterized primarily by the Stroop task rather than the flanker task supports our hypothesis; namely, that the Stroop task involves greater interference suppression at the response level due to spatial-response conflict, making it more sensitive to the motor pathology of PD. Consequently, combining measures of interference suppression and recognition memory deficits appears to be the most effective strategy for detecting cognitive impairment in patients with PD.

We acknowledge that our study has several limitations that should be addressed in future research. First, we lacked neuroimaging data, which could have helped identify the underlying neural bases of the observed cognitive impairments, such as the specific nature of the memory deficits. Second, our investigation focused on specific attention and memory tasks; consequently, we did not cover other cognitive domains or employ tasks that might have been more sensitive to detecting subtle group differences. Finally, given the well-documented heterogeneity of cognitive decline in PD, it is possible that our sample consisted of patients whose specific deficits aligned with the areas detected by our analysis. Therefore, these findings may not perfectly replicate in other patient samples with different clinical profiles.

In conclusion, our results suggest that it is possible to identify cognitive deficits in PD patients without needing an extensive number of cognitive tasks, thereby avoiding the redundancy of assessing the same underlying deficits with multiple tests. Additionally, computerized and time-based measures of attention represent a valid approach to assessing cognitive deficits in combination with traditional neuropsychological assessment.

## Data Availability

All data produced in the present study are available upon reasonable request to the authors.
The faces appearing in Figure 3 where downloaded from this database:
https://faces.mpdl.mpg.de/imeji/

https://doi.org/10.17605/OSF.IO/VGHY8

## CRediT authorship contribution statement

*Marco Calabria:* Conceptualization, Methodology, Formal analysis, Investigation, Funding acquisition, Writing - Original Draft, Writing - Review & Editing

*Lucas Guallar:* Methodology, Investigation, Writing - Original Draft, Writing - Review & Editing *Carmen García-Sánchez:* Methodology, Investigation, Resources, Writing - Original Draft, Writing - Review & Editing

*Berta Pascual Sedano:* Methodology, Investigation, Resources, Writing - Original Draft, Writing - Review & Editing

*Jaime Kulisevsky:* Investigation, Resources, Writing - Original Draft, Writing - Review & Editing

## Declaration of generative AI and AI-assisted technologies in the manuscript preparation process

During the preparation of this work, the authors used Gemini for proofreading the text originally written without the assistance of AI. After using this tool/service, the authors reviewed and edited the content as needed and took full responsibility for the content of the published article.

## Acknowledgements

The authors would like to thank all the study participants for their collaboration in this study.

