## Supplementary Material for "Attention and memory in Parkinson’s disease: a discriminant analysis approach"

**APPENDIX A – Supplementary Material**

1. **Marginal means of the task performance for cognitively unimpaired and patients with PD**

|  | **CU** | | **PD** | |
| --- | --- | --- | --- | --- |
| ***Task*** | *Means* | *SE* | *Means* | *SE* |
| **Flanker task** |  |  |  |  |
| *Congruent trials* |  |  |  |  |
| Accuracy | 0.99 | 0.01 | 0.99 | 0.01 |
| RTs | 725 | 25.5 | 737 | 37.1 |
| *Incongruent trials* |  |  |  |  |
| Accuracy | 0.98 | 0.01 | 0.96 | 0.02 |
| RTs | 845 | 29.8 | 889 | 45.3 |
| **Spatial Stroop** |  |  |  |  |
| *Congruent trials* |  |  |  |  |
| Accuracy | 0.99 | 0.01 | 0.98 | 0.01 |
| RTs | 534 | 24.2 | 550 | 21.9 |
| *Incongruent trials* |  |  |  |  |
| Accuracy | 0.99 | 0.01 | 0.92 | 0.02 |
| RTs | 601 | 27.2 | 645 | 25.7 |
| **Memory task** |  |  |  |  |
| d' values | 1.35 | 0.20 | 0.27 | 0.17 |
| c values | 0.26 | 0.13 | 0.40 | 0.11 |
| **n-back task** |  |  |  |  |
| d' values | 1.62 | 0.25 | 0.94 | 0.22 |
| c values | 0.21 | 0.12 | 0.12 | 0.11 |

2. **Flanker task: Accuracy**

**Model:** Accuracy ~ CONGRUENCY * GROUP + MoCA + Age + Education + (1 | ID) + (1 | ITEM)

|  | **ACCURACY** | | |
| --- | --- | --- | --- |
| *Predictors* | *Log-Odds* | *CI* | *p* |
| (Intercept) | 5.62 | 4.87 – 6.37 | **<0.001** |
| CONGRUENCY [Incongruent] | -1.69 | -2.43 – -0.96 | **<0.001** |
| GROUP [PD] | -0.36 | -1.68 – 0.96 | 0.594 |
| MoCA | 0.57 | 0.07 – 1.07 | **0.025** |
| Age | -0.16 | -0.45 – 0.13 | 0.274 |
| Education | -0.20 | -0.45 – 0.05 | 0.120 |
| CONGRUENCY [Incongruent] × GROUP [PD] | -0.44 | -1.37 – 0.49 | 0.351 |
| **Random Effects** | | | |
| σ^2^ | 3.29 | | |
| τ_00_ _ID_ | 0.60 | | |
| τ_00_ _ITEM_ | 0.17 | | |
| ICC | 0.19 | | |
| N _ID_ | 55 | | |
| N _ITEM_ | 33 | | |
| Observations | 7200 | | |
| Marginal R^2^ / Conditional R^2^ | 0.268 0.408 | | |

3. **Flanker task: RTs**

**Model:** log(RT) ~ CONGRUENCY * GROUP + MoCA+ Age + Education + (1 | ID) + (1 | ITEM)

|  | **log(RT)** | | |
| --- | --- | --- | --- |
| *Predictors* | *Estimates* | *CI* | *p* |
| (Intercept) | 6.59 | 6.52 – 6.65 | **<0.001** |
| CONGRUENCY [Incongruent] | 0.15 | 0.12 – 0.19 | **<0.001** |
| GROUP [PD] | 0.02 | -0.12 – 0.16 | 0.813 |
| MoCA | -0.02 | -0.08 – 0.04 | 0.549 |
| Age | 0.01 | -0.03 – 0.04 | 0.673 |
| Education | -0.03 | -0.07 – 0.00 | 0.056 |
| CONGRUENCY [Incongruent] × GROUP [PD] | 0.03 | 0.01 – 0.06 | **0.014** |
| **Random Effects** | | | |
| σ^2^ | 0.03 | | |
| τ_00_ _ID_ | 0.02 | | |
| τ_00_ _ITEM_ | 0.00 | | |
| ICC | 0.41 | | |
| N _ID_ | 55 | | |
| N _ITEM_ | 33 | | |
| Observations | 6816 | | |
| Marginal R^2^ / Conditional R^2^ | 0.134 / 0.493 | | |

4. **Stroop task: Accuracy**

**Model:** Accuracy ~ CONGRUENCY * GROUP + MoCA + Age + Education + (1 | ID) + (1 | ITEM)

|  | **ACCURACY** | | |
| --- | --- | --- | --- |
| *Predictors* | *Log-Odds* | *CI* | *p* |
| (Intercept) | 5.97 | 4.72 – 7.23 | **<0.001** |
| CONGRUENCY [Incongruent] | -1.28 | -2.37 – -0.19 | **0.022** |
| GROUP [PD] | -1.52 | -3.17 – 0.13 | 0.071 |
| MoCA z | -0.09 | -0.76 – 0.58 | 0.792 |
| Age z | -0.23 | -0.63 – 0.18 | 0.274 |
| Education z | -0.08 | -0.44 – 0.28 | 0.655 |
| CONGRUENCY [Incongruent]× GROUP [PD] | -0.11 | -1.18 – 0.96 | 0.841 |
| **Random Effects** | | | |
| σ^2^ | 3.29 | | |
| τ_00_ _ID_ | 1.08 | | |
| τ_00_ _ITEM_ | 0.10 | | |
| ICC | 0.26 | | |
| N _ID_ | 55 | | |
| N _ITEM_ | 8 | | |
| Observations | 5438 | | |
| Marginal R^2^ / Conditional R^2^ | 0.180 / 0.396 | | |

5. **Stroop task: RTs**

**Model:** log(RT) ~ CONGRUENCY * GROUP + MoCA + Age + Education + (1 | ID) + (1 | ITEM)

|  | **log(RT)** | | |
| --- | --- | --- | --- |
| *Predictors* | *Estimates* | *CI* | *p* |
| (Intercept) | 6.28 | 6.19 – 6.37 | **<0.001** |
| CONGRUENCY [Incongruent] | 0.12 | 0.09 – 0.15 | **<0.001** |
| GROUP [PD] | 0.03 | -0.11 – 0.17 | 0.691 |
| MoCA | -0.05 | -0.12 – 0.02 | 0.200 |
| Age | -0.01 | -0.05 – 0.03 | 0.670 |
| Education | -0.03 | -0.06 – 0.01 | 0.161 |
| CONGRUENCY [Incongruent] × GROUP [PD] | 0.04 | 0.02 – 0.06 | **<0.001** |
| **Random Effects** | | | |
| σ^2^ | 0.04 | | |
| τ_00_ _ID_ | 0.02 | | |
| τ_00_ _ITEM_ | 0.00 | | |
| ICC | 0.32 | | |
| N _ID_ | 55 | | |
| N _ITEM_ | 8 | | |
| Observations | 5192 | | |
| Marginal R^2^ / Conditional R^2^ | 0.139 / 0.412 | | |

6. **Recognition memory task**

**Model:** D’ ~ GROUP + Age+ Education + MoCA

|  | **D’ values** | | |
| --- | --- | --- | --- |
| *Predictors* | *Estimates* | *CI* | *p* |
| (Intercept) | 1.35 | 0.95 – 1.75 | **<0.001** |
| GROUP [PD] | -1.08 | -1.75 – -0.41 | **0.002** |
| Age | -0.15 | -0.35 – 0.04 | 0.120 |
| Education | 0.05 | -0.12 – 0.23 | 0.539 |
| MoCA | 0.06 | -0.27 – 0.39 | 0.717 |
| Observations | 55 | | |
| R^2^ / R^2^ adjusted | 0.494 / 0.454 | | |

**Model:** C ~ GROUP + Age+ Education + MoCA

|  | **C values** | | |
| --- | --- | --- | --- |
| *Predictors* | *Estimates* | *CI* | *p* |
| (Intercept) | 0.26 | 0.00 – 0.51 | **0.050** |
| GROUP [PD] | 0.14 | -0.29 – 0.57 | 0.508 |
| Age | -0.13 | -0.26 – -0.01 | **0.035** |
| Education | -0.14 | -0.26 – -0.03 | **0.014** |
| MoCA | 0.02 | -0.19 – 0.23 | 0.842 |
| Observations | 55 | | |
| R^2^ / R^2^ adjusted | 0.224 / 0.162 | | |

7. **N-back task**

**Model:** D’ ~ GROUP + Age+ Education + MoCA

|  | **D’ values** | | |
| --- | --- | --- | --- |
| *Predictors* | *Estimates* | *CI* | *p* |
| (Intercept) | 1.62 | 1.12 – 2.13 | **<0.001** |
| Group [PD] | -0.68 | -1.53 – 0.16 | 0.109 |
| Age | -0.29 | -0.53 – -0.04 | **0.023** |
| Education | 0.21 | -0.01 – 0.43 | 0.062 |
| MoCA | -0.04 | -0.45 – 0.37 | 0.835 |
| Observations | 55 | | |
| R^2^ / R^2^ adjusted | 0.225 / 0.163 | | |

**Model:** C ~ GROUP + Age+ Education + MoCA

|  | **C values** | | |
| --- | --- | --- | --- |
| *Predictors* | *Estimates* | *CI* | *p* |
| (Intercept) | 0.21 | -0.04 – 0.46 | 0.099 |
| Group [PD] | -0.09 | -0.51 – 0.33 | 0.663 |
| Age z | 0.01 | -0.11 – 0.14 | 0.829 |
| Education z | -0.10 | -0.21 – 0.01 | 0.083 |
| MoCA z | 0.07 | -0.14 – 0.27 | 0.512 |
| Observations | 55 | | |
| R^2^ / R^2^ adjusted | 0.149 / 0.081 | | |
